# Rising resistance to linezolid among critically ill patients calls for model-informed precision dosing in the empirical setting

**DOI:** 10.64898/2025.11.28.25340407

**Authors:** Johannes Starp, Antonia Leonhardt, Jette Jung, Maximilian Sachsenberg Stoschus, Michael Zoller, Christina Scharf, Johannes Zander, Michael Paal, Sebastian G. Wicha, Uwe Liebchen

## Abstract

Linezolid is important for treating infections caused by Gram-positive bacteria resistant to first-line agents. Severe infections often require empirical therapy before pathogen identification and susceptibility testing. This evaluation aims to compare different empirical linezolid dosing strategies in intensive care unit (ICU) patients to improve pathogen eradication while minimising toxicities. Three dosing strategies were compared: (i) standard dosing, (ii) therapeutic drug monitoring (TDM) guided dosing, and (iii) a theoretical model-informed precision dosing (MIPD) strategy based on Bayesian forecasting with a population pharmacokinetic model. Each scenario was combined with an ICU-specific minimal inhibitory concentration (MIC) distribution comprising more than 19,000 isolates collected over 10 years. Data from 117 critically ill patients with 2,184 TDM samples were available. Linezolid resistance was highest in ICU isolates (9.6%) compared with isolates from the intermediate care unit (4.3%), general wards (3.6%), and emergency room (1.6%). Among ICU isolates, resistance increased significantly over the 10-year study period, from 6.3% in 2015 to 11.4% in 2024. Empirical target attainment was 36.6% with standard dosing and 48.5% with TDM-guided dose adaptation, whereas theoretical MIPD projected target attainment in 71.4-81.6% of patients. After excluding resistant isolates, the corresponding values were 40.5%, 53.7%, and 79.0-90.3%, respectively. In the era of increasing antimicrobial resistance, novel dosing strategies such as MIPD become urgently necessary. Our results suggest MIPD may represent a promising strategy improving target attainment compared to standard regimens and TDM-guided dosing which frequently fail to achieve adequate target exposure in ICU patients.

## 1 Introduction

Antimicrobial resistance continues to limit therapeutic options and increases the need for more efficient use of existing antibiotics. Hence, the pivotal question is how to individualise treatment to administer the right antibiotic and the right dose for each patient.

Linezolid is an oxazolidinone antibiotic used to treat infections with Gram-positive pathogens, particularly when first-line therapies fail against methicillin-resistant *Staphylococcus aureus* (MRSA) or vancomycin-resistant enterococci (VRE). The standard intravenous regimen of 600 mg twice daily is commonly administered without initial dose adjustment. However, linezolid pharmacokinetics vary substantially between patients, especially in ICU patients, and retrospective analyses indicate that approximately half of the patients receiving standard dosing are either underexposed or overexposed (Pea et al. 2017). Overexposure is associated with haematological toxicity, such as thrombocytopenia, sometimes necessitating dose reduction or discontinuation (Nukui et al. 2013; Dong et al. 2014; Matsumoto et al. 2014). Conversely, underexposure may compromise antibacterial efficacy and increase the risk of treatment failure.

Linezolid efficacy is commonly assessed by pharmacokinetic-pharmacodynamic (PK/PD) parameters such as the percentage of time above minimal inhibitory concentration (MIC) (82-99%t > MIC) or the ratio of the area under the concentration-time curve to MIC (AUC/MIC). The pharmacological adequacy of linezolid therapy cannot be assessed from drug concentrations alone, because antibacterial activity also depends on the susceptibility of the infecting pathogen. This relationship is described by pharmacokinetic/pharmacodynamic (PK/PD) indices that relate systematic exposure to the minimum inhibitory concentration (MIC), including the percentage of time that concentrations remain above the MIC (82-99%t > MIC) and the ratio of the area under the concentration-time curve to the MIC (AUC/MIC > 85-164 mg/L*h) (Wicha et al. 2021). For a given exposure, the probability of achieving an efficacy target decreases as the MIC increases. Accordingly, PK/PD target attainment represents the probability that a dosing regimen provides exposure expected to support antibacterial activity against a pathogen with a specific MIC.

In targeted therapy, PK/PD target attainment can be assessed using the MIC of the identified pathogen. During empirical therapy, however, treatment must be initiated before the causative pathogen and its susceptibility are known. Microbiological results may not become available until several days after sample collection, MIC values are not routinely reported by all laboratories, and individual MIC measurements are subject to methodological uncertainty (Mouton et al. 2018; Liebchen et al. 2020). When no pathogen-specific MIC is available, the expected performance of a dosing regimen can instead be estimated by combining its probability of target attainment at each MIC with the frequency of these MIC values in a representative local susceptibility distribution. This approach integrates variability in both patient pharmacokinetics and pathogen susceptibility and therefore provides a pharmacological framework for evaluating empirical dosing.

Therapeutic drug monitoring (TDM) can support dose individualisation by relating measured concentrations to predefined exposure targets. Population pharmacokinetic models additionally describe expected concentration–time profiles and sources of interindividual variability. Combining these models with individual patient characteristics and measured concentrations through Bayesian forecasting forms the basis of model-informed precision dosing (MIPD) (Minichmayr et al. 2024). Numerous popPK models for linezolid have been published and in a recent multicentre evaluation study we identified candidate models adequately predicting pharmacokinetics of critically ill patients (Starp et al. 2026). In contrast to the elaboration on pharmacokinetics in recent years, limited information is available on contemporary MIC distributions in ICU populations (Whitehouse et al. 2005; Roger et al. 2016; Minichmayr et al. 2017; Taubert et al. 2017; Cojutti et al. 2019; Ehmann et al. 2020; Milaković et al. 2024).

This study aimed to optimise empirical linezolid dosing in a setting where neither the causative pathogen nor its MIC is yet known, drawing on a large ICU-specific MIC database. Empirical target attainment was evaluated for (i) standard dosing, (ii) TDM-guided dosing, and (iii) MIPD-based dosing. This combined approach sought to identify clinical scenarios with potential for dosing optimisation in this vulnerable population and to highlight promising, individualised dosing strategies.

## 2 Methods

### 2.1 MIC distribution and resistance trends

The distribution of MICs of Gram-positive pathogens isolated from adult patients at LMU University Hospital, Munich, between 2015 and 2024 was retrospectively collected across different hospital wards, including the ICU, emergency room, intermediate care unit, and general wards (Fig. 1). However, the present evaluation of empirical target attainment focused exclusively on ICU data.

**Fig. 1.**
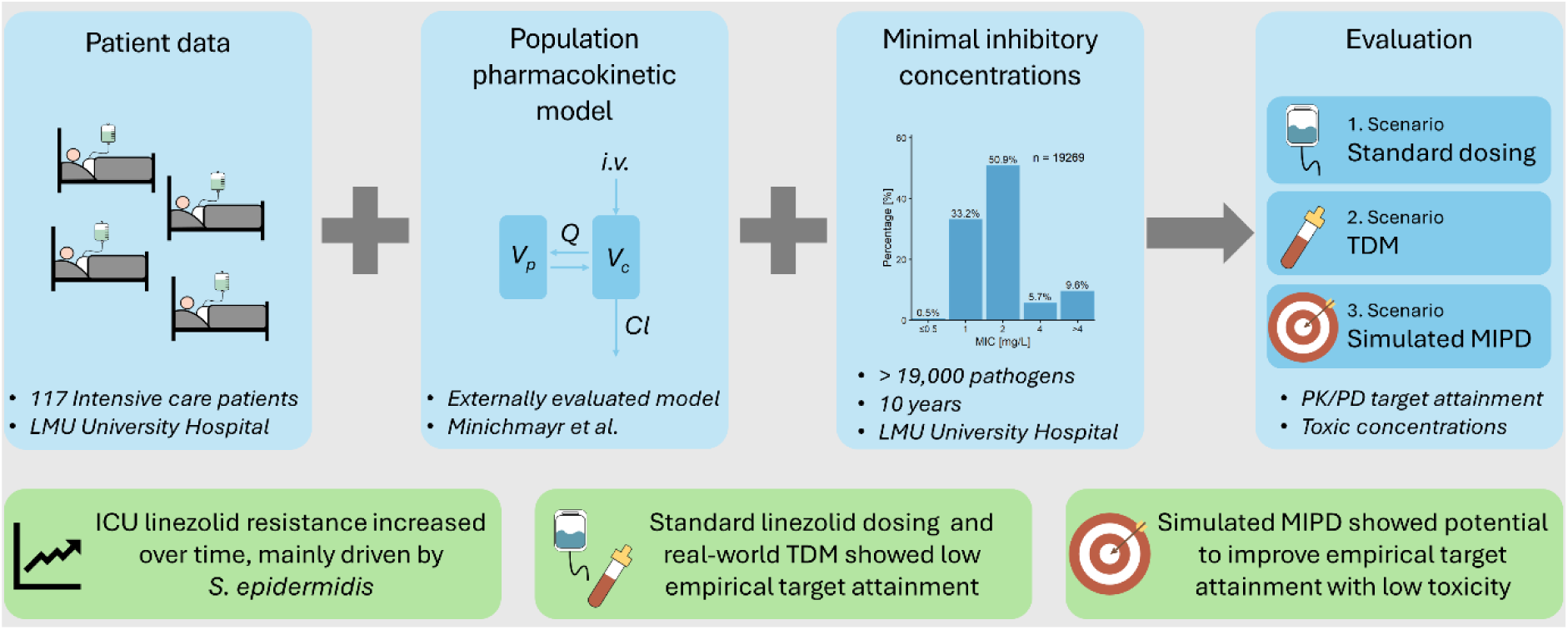
Graphical abstract. Cl, clearance; i.v., intravenously; MIPD, model-informed precision dosing, Q, intercompartimental clearance; TDM, therapeutic drug monitoring; Vc, central volume of distribution; Vp, peripheral volume of distribution

Susceptibility testing was conducted using a BD Phoenix Automated Microbiology System and according to the manufacturer’s protocol. Fresh colonies were suspended in 4.5 mL BD Phoenix ID broth, and the turbidity was adjusted to 0.5 McFarland with a BD PhoenixSpec nephelometer. A 25 µL aliquot of the standardized suspension was added to BD Phoenix broth, followed by a single drop of the colorimetric redox indicator. The inoculated medium was dispensed into an PMIC-88 panel containing linezolid at 0.5– 4 µg/mL in two-fold dilutions, then incubated in a BD Phoenix M50 instrument. Minimum inhibitory concentration (MIC) values were subsequently extracted directly from the BD EpiCenter software. To ensure unique data points, the dataset was filtered to exclude duplicate isolates from the same pathogen, patient, and sampling material. Resistance was defined as MIC > 4 mg/L in accordance to the clinical breakpoints for enterococci and MRSA (EUCAST 2026). To evaluate temporal trends, yearly percentages of resistant isolates were assessed using linear regression. Statistical significance of the slope was evaluated with a two-sided t-test.

### 2.2 Patient data

Data from two previously published prospective observational studies conducted at LMU University Hospital were analysed: study 1 (ClinicalTrials.gov NCT01793012; registered 24 January 2013) and study 2 (ClinicalTrails.gov NCT03985605; registered 14 June 2019) (Zoller et al. 2014; Taubert et al. 2016; Zoller et al. 2022). In both studies, linezolid therapy was initiated at the standard dose of 600 mg twice daily administered as a short infusion. Throughout therapy, dosing either remained as standard treatment in study 1 or was individualised in study 2 based on TDM and physician discretion.

### 2.3 Pharmacokinetic modelling and evaluated scenarios

The previously published popPK model by Minichmayr et al. (2017) was used based on an external evaluation demonstrating its suitability for *a posteriori* (Bayesian) predictions (Starp et al. 2026). The model code is provided in the supplementary material (1.1). Individual predictions were calculated using maximum *a posteriori* Bayesian estimates obtained with the first-order conditional estimation with interaction (FOCEI) method (MAXEVAL=0). The analyses were performed using NONMEM® (version 7.5) in conjunction with Pirana (version 23.10.1) and PsN (version 5.6.0). Data management, statistical analyses, and graphical representations were conducted using R (version 4.5.1) and RStudio® (version 2026.06.0).

This study compared three scenarios: (i) standard dosing, (ii) TDM-guided dosing and (iii) a theoretical MIPD scenario. For comparison across strategies, observed dosing intervals (ODI) were defined as the interval between two dosing events during which at least one TDM sample was obtained. All scenarios were evaluated in the third ODI (ODI3), ensuring that at least two TDM samples were available to refine dose adjustments in the (ii) TDM and (iii) MIPD scenarios.

Because TDM samples were not consistently collected at trough time points, minimal concentrations (C_min_) were calculated using maximum *a posteriori* estimation. Individual predictions for both the (i) standard dosing and the (ii) TDM strategy were informed by all available TDM samples, with C_min_ defined as the predicted concentration at the dosing event following ODI3.

The (iii) MIPD scenario utilised both datasets and evaluated a Bayesian setting in ODI3. The calculation of individual predictions was informed by one TDM sample from ODI1 and one from ODI2. In this scenario, trough concentrations in ODI3 were approximated by the expected C_min_ (eC_min_), which were calculated by scaling the target concentration for C_min_ (4 mg/L), by the ratio of the individual prediction (C_pred_) to the actual measured concentration (C_obs_) as shown in equation 1.

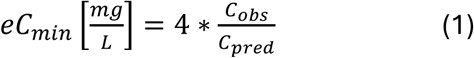

To establish a baseline across both datasets, C_min_ was calculated for the first dosing interval, during which all patients in both studies received standard dosing. The baseline scenario was informed by all TDM samples. All scenarios are summarised in Tab. 1.

**Tab. 1.**
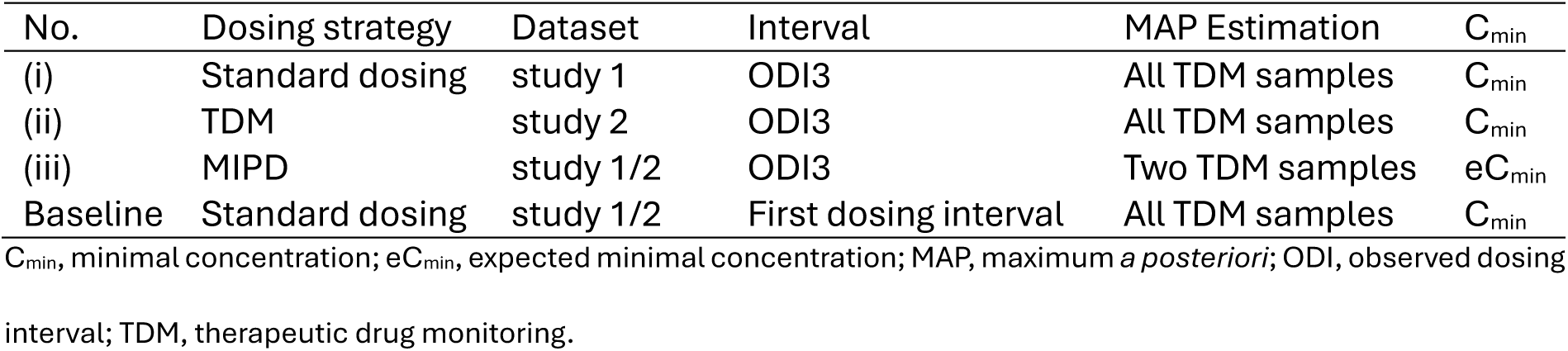
Overview of the evaluated scenarios.

| No. | Dosing strategy | Dataset | Interval | MAP Estimation | $C_{\min}$ |
| --- | --- | --- | --- | --- | --- |
| (i) | Standard dosing | study 1 | ODI3 | All TDM samples | $C_{\min}$ |
| (ii) | TDM | study 2 | ODI3 | All TDM samples | $C_{\min}$ |
| (iii) | MIPD | study 1/2 | ODI3 | Two TDM samples | $eC_{\min}$ |
| Baseline | Standard dosing | study 1/2 | First dosing interval | All TDM samples | $C_{\min}$ |

$C_{\min}$ , minimal concentration; $eC_{\min}$ , expected minimal concentration; MAP, maximum *a posteriori*; ODI, observed dosing interval; TDM, therapeutic drug monitoring.

### 2.4 PK/PD targets and toxicity thresholds

The efficacy target was defined as a minimum concentration (C_min_) above the minimum inhibitory concentration (MIC), whereas C_min_ values at or below the MIC were considered subtherapeutic. Toxicity risk was assessed using a C_min_ threshold of 8 mg/L, above which an increased risk of toxicity was assumed (Nukui et al. 2013; Dong et al. 2014; Matsumoto et al. 2014).

MIC categories from 0.5 to 4 mg/L in two-fold increments were considered susceptible, whereas isolates with MIC > 4 mg/L were classified as resistant. P_res_ denotes the percentage of resistant isolates in the respective MIC distribution. For each susceptible MIC category j, the probability of target attainment (PTA_j_) was defined (Equation 2) as the proportion of patients i who jointly met the efficacy target and non-toxicity criterion.

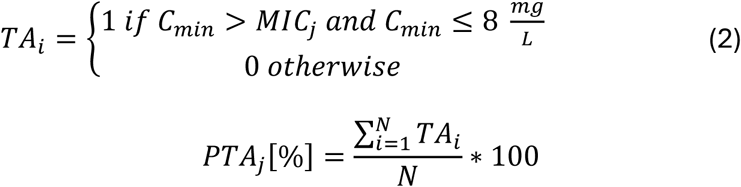

The concept of Cumulative Fraction of Response (CFR) (Mouton et al. 2005), which traditionally evaluates antibiotic efficacy for a single known pathogen, was extended to reflect the full MIC distribution of pathogens in the local setting. This allows target attainment to be assessed in the early empirical treatment phase when the causative pathogen is unknown, and corresponds to what has previously been described as the Local Pathogen Independent Fraction of Response (Weinelt et al. 2021; Liebchen et al. 2022). CFR was calculated by weighing PTA_j_ for each MIC category j by its frequency (F_j_) in the ICU-specific distribution and was referred to as Empirical target attainment (Equation 3). Resistant isolates contributed zero to the CFR.

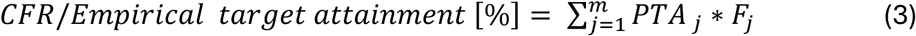

Cumulative fractions of patients at risk of toxicity (P_tox_) and those failing to achieve pathogen eradication (P_sub_) were calculated according to Equations 4 and 5:

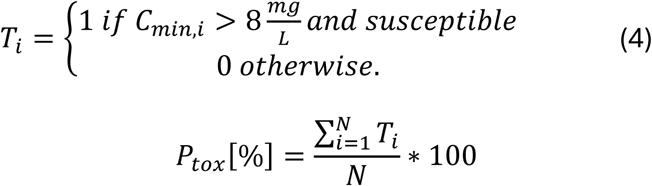

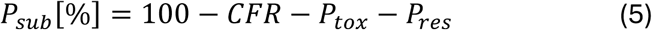

All dosing scenarios were evaluated using the ICU-specific susceptibility distribution of Gram-positive pathogens to reflect the empirical treatment setting, in which the causative pathogen is usually unknown at therapy initiation. In addition, stratified analyses were performed to account for clinical situations in which the expected pathogen spectrum may be narrowed based on the presumed site of infection. For example, pneumonia was represented by the *S. aureus* MIC distribution and abdominal infections by the enterococcal MIC distribution.

## 3 Results

### 3.1 MIC distribution and resistance trends

During the 10-year period of linezolid susceptibility testing a total of 68,089 samples were collected split in ICU (n = 19,269), intermediate care unit (n = 1,887), general ward (n = 41,353), and emergency room (n = 5,580). Resistance rates were higher in the ICU (9.6%) compared to the intermediate care unit (4.3%), general ward (3.6%), and emergency room (1.6%) (Fig. 2). Tab. 2 summarises the MIC distribution in the ICU.

**Fig. 2.**
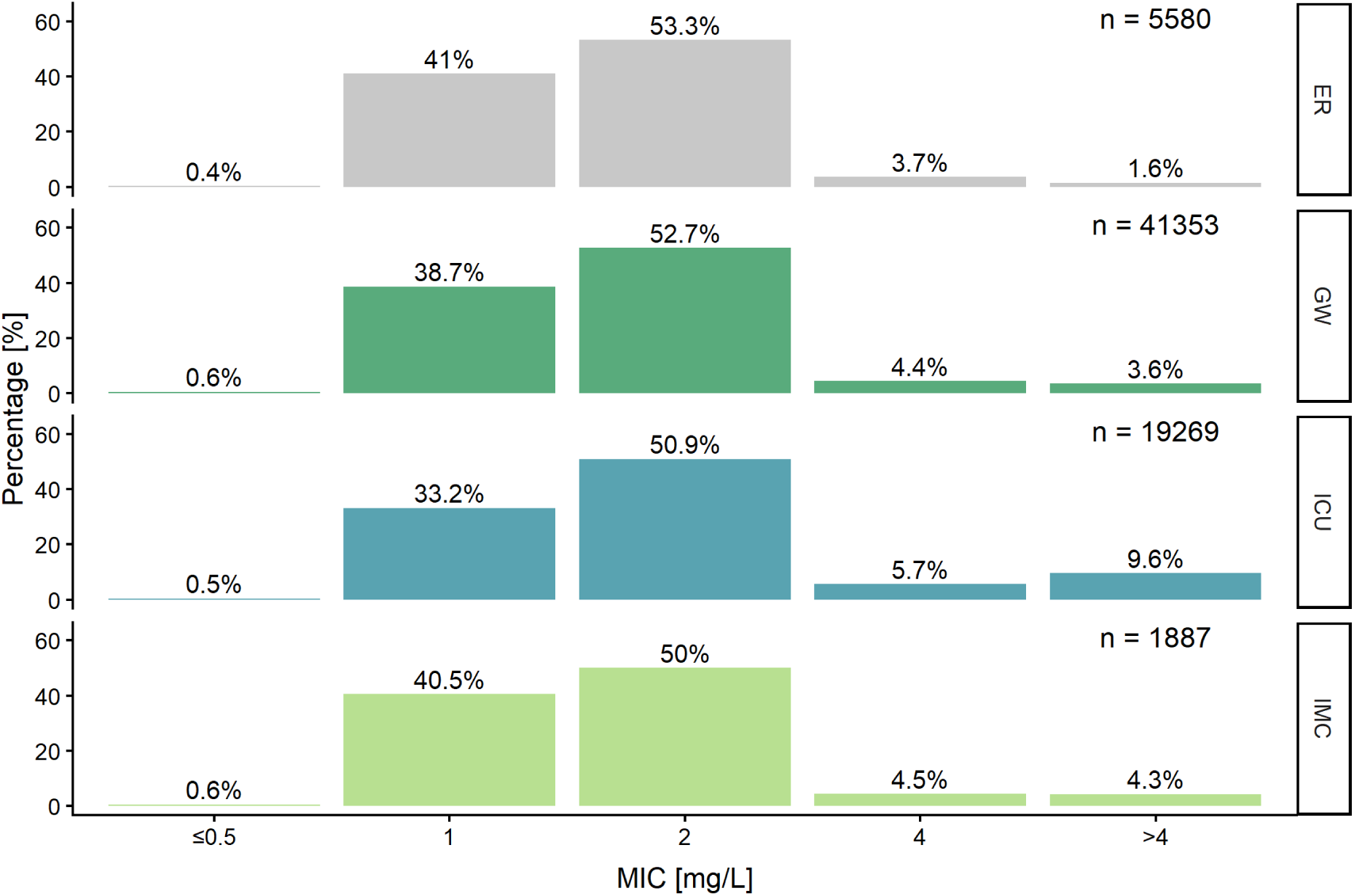
Distribution of minimal inhibitory concentration (MIC) at different wards of LMU University Hospital. ER, emergency room; GW, general ward; ICU, intensive care unit; IMC, intermediate care unit

**Tab. 2.** Distribution of minimal inhibitory concentrations of pathogens collected on the intensive care unit and tested for linezolid.

|  | Minimal inhibitory concentration [mg/L]<sup>a</sup> |  |  |  |  |  |
| --- | --- | --- | --- | --- | --- | --- |
|  | n | ≤0.5 | 1 | 2 | 4 | >4 |
| Intensive care unit | 19269 | 94 (0.5) | 6399 (33.2) | 9815 (50.9) | 1105 (5.7) | 1856 (9.6) |
| Year |  |  |  |  |  |  |
| 2015 | 1563 | 9 (0.6) | 514 (32.9) | 785 (50.2) | 157 (10) | 98 (6.3) |
| 2016 | 2009 | 5 (0.2) | 743 (37) | 1012 (50.4) | 118 (5.9) | 131 (6.5) |
| 2017 | 1936 | 4 (0.2) | 546 (28.2) | 1145 (59.1) | 131 (6.8) | 110 (5.7) |
| 2018 | 1837 | 3 (0.2) | 487 (26.5) | 1117 (60.8) | 90 (4.9) | 140 (7.6) |
| 2019 | 2150 | 6 (0.3) | 579 (26.9) | 1290 (60) | 108 (5) | 167 (7.8) |
| 2020 | 2313 | 9 (0.4) | 583 (25.2) | 1366 (59.1) | 129 (5.6) | 226 (9.8) |
| 2021 | 2133 | 10 (0.5) | 709 (33.2) | 1061 (49.7) | 84 (3.9) | 269 (12.6) |
| 2022 | 1898 | 19 (1) | 812 (42.8) | 740 (39) | 87 (4.6) | 240 (12.6) |
| 2023 | 2349 | 16 (0.7) | 954 (40.6) | 894 (38.1) | 133 (5.7) | 352 (15) |
| 2024 | 1081 | 13 (1.2) | 472 (43.7) | 405 (37.5) | 68 (6.3) | 123 (11.4) |
| Pathogen |  |  |  |  |  |  |
| Enterococcus faecalis | 2339 | 39 (1.7) | 1803 (77.1) | 468 (20) | 10 (0.4) | 19 (0.8) |
| Enterococcus faecium | 5845 | 13 (0.2) | 1006 (17.2) | 3924 (67.1) | 587 (10) | 315 (5.4) |
| Enterococcus, other spp. | 81 | 1 (1.2) | 10 (12.3) | 36 (44.4) | 19 (23.5) | 15 (18.5) |
| Staphylococcus aureus | 3658 | 6 (0.2) | 1158 (31.7) | 2408 (65.8) | 55 (1.5) | 31 (0.8) |
| Staphylococcus epidermidis | 5596 | 26 (0.5) | 1225 (21.9) | 2514 (44.9) | 405 (7.2) | 1426 (25.5) |
| Staphylococcus, other spp. | 1741 | 9 (0.5) | 1197 (68.8) | 465 (26.7) | 29 (1.7) | 41 (2.4) |
| Unspecified pathogen | 9 | - | - | - | - | 9 (100) |

<sup>a</sup>n (%)

A significant increase in the proportion of resistant isolates was observed in the ICU, rising from 6.3% in 2015 to 11.4% in 2024. Most resistant isolates (76.8%) were *S. epidermidis*, which was the primary driver of this trend, increasing significantly from 12.6% in 2015 to 33.2% in 2024. No statistically significant temporal changes were observed for the other genera shown in Fig. 3.

**Fig. 3.**
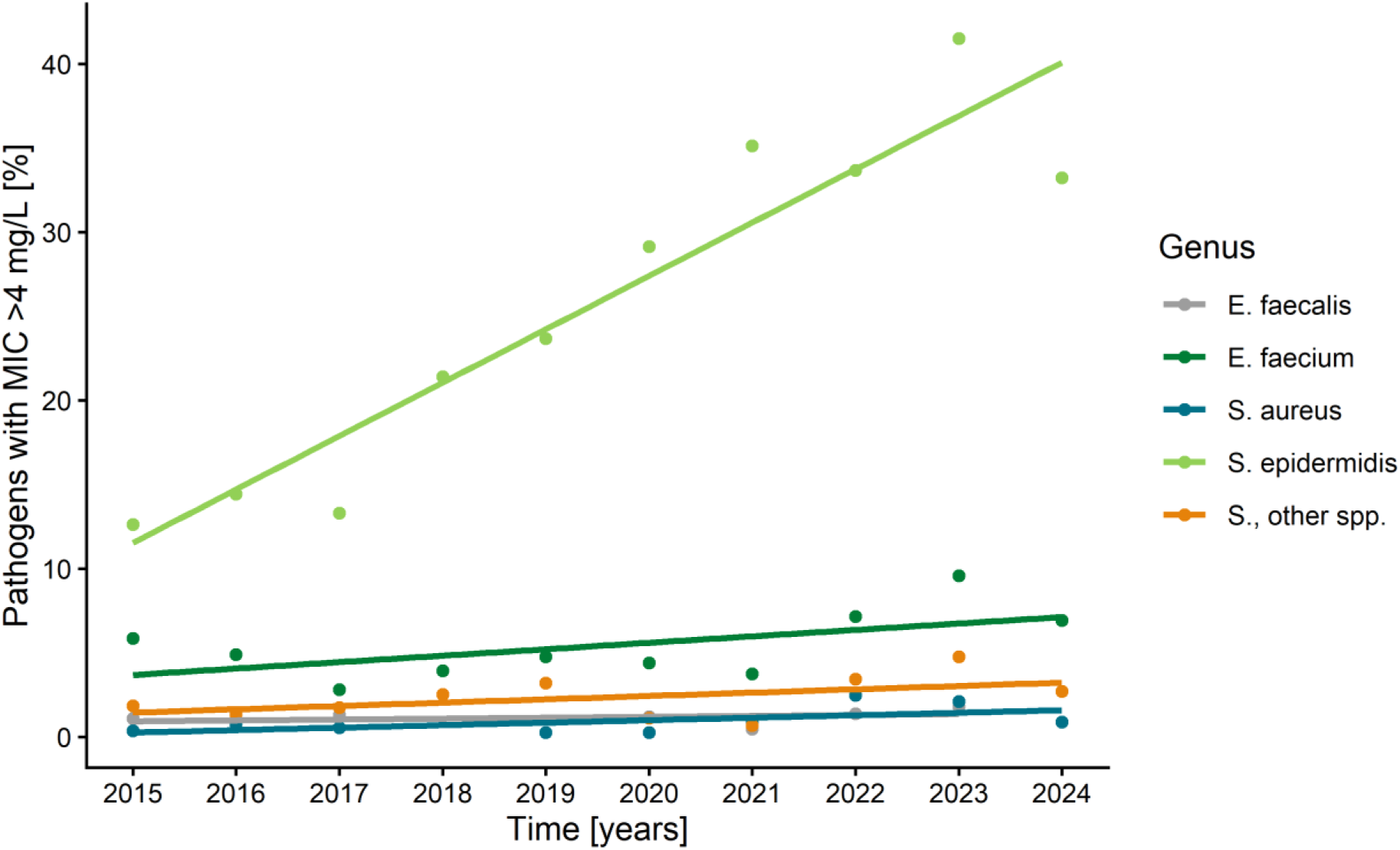
Percentage of linezolid resistant pathogens (MIC > 4 mg/L) of that genus in different years. Other subspecies of Enterococci were not listed, because overall sample size was below 100. MIC, Minimal inhibitory concentration. E., Enterococcus; S., Staphylococcus

### 3.2 Patient data

A total of 117 patients were included for analysis (Tab. 3). In comparison to the originally published datasets no patient was excluded in the study 1 dataset, whereas three patients were excluded from the study 2 dataset due to implausible height record (n = 1), less than three TDM samples (n = 1), and therapy duration shorter than 48 h (n = 1). In total, 2184 TDM samples were available across the treatment courses. The timing of sample collection relative to the first and previous doses is shown in supplementary Tab. A1.

**Tab. 3.** Patients’ characteristics at day one of therapy (Zoller et al. 2014, 2022; Taubert et al. 201c)

| Characteristic | Overall<sup>a</sup> (n = 117) | Study 1<sup>a</sup> (n = 52) | Study 2<sup>a</sup> (n = 65) |
| --- | --- | --- | --- |
| Female<sup>b</sup> | 36 (31%) | 19 (37%) | 17 (26%) |
| Male<sup>b</sup> | 81 (69%) | 33 (63%) | 48 (74%) |
| Age [years] | 59 [22-87] | 58 [22-84] | 61 [23-87] |
| Weight [kg] | 74 [40-170] | 75 [44-120] | 74 [40-170] |
| Height [cm] | 174 [150-196] | 172 [150-196] | 175 [156-192] |
| Body-mass index [kg/m<sup>2</sup>] | 24.7 [15.6-53.7] | 25.6 [15.6-37.0] | 24.7 [16.4-53.7] |
| Samples total | 2184 | 1654 | 530 |
| Samples per patient | 15 [3-51] | 32 [18-51] | 7 [3-30] |
| Samples per ODI<sup>c</sup> | 1 [1-9.5] | 5.75 [3.5-9.5] | 1 [1-2] |
| Creatinine [mg/dL] | 1.20 [0.30-6.70] | 1.15 [0.40-2.80] | 1.20 [0.30-6.70] |
| Creatinine clearance (Cockcroft-Gault) [mL/min] | 66 [10-323] | 74 [19-172] | 64 [10-323] |
| Platelets [10<sup>3</sup>/μL] | 152 [4-941] | 167 [32-577] | 139 [4-941] |
| White Blood Cells [10<sup>9</sup>/L] | 11 [0-42] | 13 [5-36] | 9 [0-42] |
| Albumin [g/L] | 26.0 [18.0-43.0] | 27.0 [19.0-43.0] | 24.0 [18.0-38.0] |
| CRP [mg/L] | 11 [1-38] | 13 [1-38] | 10 [1-36] |
| APACHE | 29 [9-45] | 29 [9-45] | NA |
| ARDS | 30 (26%) | 15 (29%) | 15 (23%) |
| ECMO | 15 (13%) | 11 (21%) | 4 (6.2%) |
| RRT | 24 (21%) | 13 (25%) | 11 (17%) |

<sup>a</sup>n (%); Median [Minimum-Maximum]<sup>b</sup>Sex was recorded as binary (female or male) based on the information provided by the enrolling physician at the time of hospital admission or study enrolment. The study investigators did not redefine or reassign sex for the purposes of this analysis (WSM 1.4).<sup>c</sup>For each patient, the median number of samples per observed dosing interval (ODI) was calculated; values are reported as the median [range] of these patient-specific medians.
APACHE II, Acute physiology and chronic health evaluation; ECMO, Extracorporeal membrane oxygenation; NA, not reported; ODI, observed dosing interval; RRT, Renal replacement therapy.

### 3.3 Standard and TDM-guided dosing

During the first dosing interval, empirical target attainment with standard dosing was higher in study 2 than in study 1 (59.4% versus 35.9%; Fig. 4). When resistant isolates were excluded, target attainment among susceptible isolates was 65.7% and 39.7%, respectively.

**Fig. 4.**
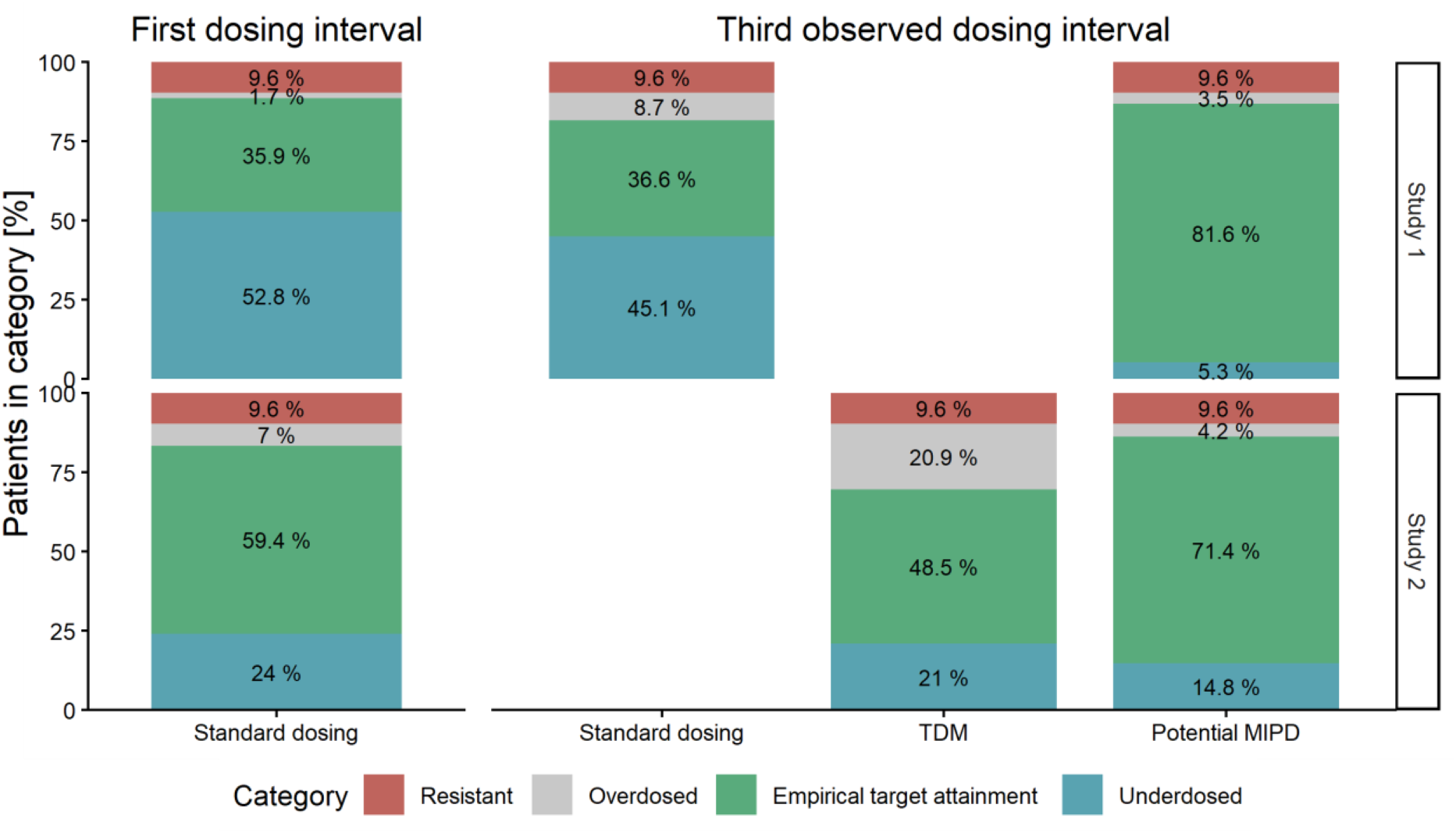
Weighted percentages of empirical outcomes during linezolid therapy under different dosing scenarios. The analysis combined patient-specific exposure with the ICU-specific MIC distribution. Outcomes were assigned to four mutually exclusive categories: resistant isolates (MIC > 4 mg/L), underdosing among susceptible isolates (Cmin ≤ MIC), empirical target attainment (MIC < Cmin ≤ 8 mg/L), and overdosing among susceptible isolates (Cmin > 8 mg/L). Standard dosing and therapeutic drug monitoring (TDM) represent observed clinical scenarios, whereas model-informed precision dosing (MIPD) represents a theoretical strategy

In study 1, empirical target attainment under continued standard dosing was 36.6% in the third observed dosing interval (ODI3), corresponding to 40.5% after exclusion of resistant isolates. Of the overall empirical outcome distribution, 45.1% was classified as underdosed, 8.7% as overdosed, and 9.6% as resistant.

In study 2, most patients continued to receive the standard regimen during the first three observed dosing intervals (ODI1: 91%, ODI2: 95%, and ODI3: 92%). The exact dosing regimens are shown in Supplementary Fig. A1. At ODI3, empirical target attainment with TDM-guided dosing was 48.5%, corresponding to 53.7% among susceptible isolates. The proportions classified as underdosed and overdosed were 21.0% and 20.9%, respectively, while 9.6% represented resistant isolates.

Compared with the first dosing interval in study 2, exposure-related underdosing decreased from 24.0% to 21.0%, whereas overdosing increased from 7.0% to 20.9%. When the alternative toxicity threshold of 10 mg/L used in the original study was applied, 3.1% and 15.4% of patients had predicted Cmin values above this threshold in ODI1 and ODI3, respectively.

### 3.4 Theoretical MIPD approach

In the theoretical MIPD scenario, projected empirical target attainment was higher than under standard or observed TDM-guided dosing, reaching 81.6% in study 1 and 71.4% in study 2. After excluding resistant isolates, projected target attainment increased to 90.3% and 79.0%, respectively. Exposure-related underdosing accounted for 5.3% in study 1 and 14.8% in study 2, while the respective proportions classified as overdosed were 3.5% and 4.2%. The remaining 9.6% represented resistant isolates for which target attainment was considered unachievable irrespective of dose adaptation.

When the evaluation was stratified according to pathogen-specific ICU MIC distributions, the projected proportion of underdosed patients for the MIPD approach was lower for enterococci (6.4% in study 1 and 16.5% in study 2) and *S. aureus* (3.4% in study 1 and 15.1% in study 2), as shown in supplementary Tab. A2.

## 4 Discussion

Investigating three different empirical dosing strategies, our evaluation indicates that standard and TDM guided linezolid dosing, as implemented in the analysed cohorts, provide insufficient empirical target attainment for ICU patients, given a representative MIC distribution for Gram-positive pathogens. Implementing TDM, followed by systematic dose adjustments in a MIPD setting, could help achieving both efficacy and safety in critically ill populations.

This analysis revealed considerable variability in baseline target attainment (35.9% vs. 59.4%), underscoring the heterogeneity of critically ill patients. The differing patient collectives likely contributed to these variations: study 1 included a higher proportion of patients with pulmonary infections (67.3%), who are reported to be more prone to subtherapeutic C_min_ (Zoller et al. 2022), explaining the larger number of underdosed patients in the first (52.8%) and third (45.1%) dosing intervals. In contrast, study 2 had fewer pulmonary infections (44.8%), resulting in fewer underdosed patients (21.0– 24.0%), while the non-pulmonary subgroup (55.2%) tended to be overdosed, particularly in later dosing intervals (20.9%). Such variability calls for individualised dosing approaches like TDM or MIPD.

Shi et al. (2023) similarly investigated the effect of pharmaceutical advice and MIPD in linezolid therapy. After initiation with standard dosing, only 15.5% of C_min_ values at 48 h were within the target range of 2–7 mg/L, which is lower than in our evaluation (35.9%). Notably, target attainment improved markedly following MIPD-based dose adjustments, from 46.2% in the control group to 82.1% in the intervention group, aligning with our findings for TDM (48.5%) and projected MIPD (71.4–81.6%) (Shi et al. 2023). Together, these data highlight the value of MIPD in accounting for pharmacokinetic variability and its potential to enhance target attainment in critically ill patients.

A strength of our work is the use of an externally evaluated popPK model for the calculation of the individual patient concentrations from two prospective studies providing high confidence in the (i) standard dosing and the (ii) TDM guided scenario (Starp et al. 2026).

Another important strength of this evaluation is the use of a comprehensive, ICU-specific MIC distribution that reflects contemporary antimicrobial resistance patterns more accurately than previous studies. While several investigations have explored linezolid dose optimisation in the context of MIC distributions (Whitehouse et al. 2005; Matsumoto et al. 2014; Roger et al. 2016; Taubert et al. 2017; Ehmann et al. 2020; Milaković et al. 2024), the present study specifically addresses the empirical treatment setting by applying a contemporary ICU-specific MIC distribution derived from more than 19,000 isolates. Most of our analyses were based on the overall ICU distribution to preserve the focus on purely empirical therapy. At the same time, the additional pathogen-stratified analyses provide clinically relevant context. When empirical therapy can already be narrowed toward likely pathogen groups, such as *S. aureus* in pneumonia or enterococci in abdominal infections, the proportion of underdosed patients was modestly lower than with the overall ICU distribution. However, underdosing remained frequent under standard and observed TDM-guided dosing, indicating that narrowing the expected pathogen spectrum alone does not remove the need for systematic dose individualisation (Tab. A2). In contrast, target attainment for *S. epidermidis* remained low even in the theoretical MIPD scenario, reflecting the unfavourable susceptibility distribution of this species and suggesting that linezolid may be a poor empirical option when *S. epidermidis* is strongly suspected.

This evaluation directly balances the risk of not achieving pathogen eradication against toxicity risk. While Taubert et al. and Ide et al. acknowledged toxicity concerns, noting that achieving a CFR of >90% placed 8-14% and 17.6-31.4% of patients at risk of toxic effects, respectively (Taubert et al. 2017; Ide et al. 2018), other comparable studies as of Whitehouse et al., Ehmann et al., Milakovic et al., or Roger et al. did not incorporate the risk of toxicities into their evaluations (Whitehouse et al. 2005; Roger et al. 2016; Ehmann et al. 2020; Milaković et al. 2024).

Considering the decrease in target attainment in the TDM setting, the low rate of dose adjustments, and the magnitude in adaptation of daily linezolid doses, it should be discussed, whether TDM was adequately reflected by study 2. In particular, samples were not consistently obtained at trough, complicating interpretation by the treating physicians and underscoring the potential value of popPK models to estimate projected C_min_. As a result, some patients’ dosage was not adapted even if TDM fell outside of target range (2– 10 mg/L) (Zoller et al. 2022), a finding that is consistent with previous observations, that reported that physicians frequently did not modify linezolid dosing despite repeated supratherapeutic concentrations (Cattaneo et al. 2016). In contrast, proactive TDM brought 47.5% of patients experiencing overexposure at least once during their treatment to the desired range in a prospective, interventional study by Cojutti et al. (2019). Accordingly, the TDM-guided arm in our study should be interpreted as a real-world TDM scenario rather than as a best-case implementation of TDM-based individualisation. This distinction is important when interpreting the theoretical MIPD scenario. The MIPD approach presented here is not intended as a direct head-to-head comparison with standard dosing or observed TDM-guided dosing. Rather, it represents a best-case, model-informed strategy designed to illustrate the potential benefit of systematic, exposure-guided dose adaptation. This scenario assumes short turnaround times, structured use of prior concentration measurements, and sufficient flexibility in dose administration. It therefore should be seen as a proof-of-concept analysis that defines a possible direction for future improvement, not as evidence that MIPD is already superior in routine practice.

A limitation of our study is the theoretical nature of the MIPD scenario, though Shi et al. (2023) yield comparable results, it needs to be confirmed in a real-world setting. Additionally, the MIC distribution comprised all pathogens tested for linezolid in the ICU; however, not all patients were treated with linezolid.

## 5 Conclusions

Our analysis highlights the challenge of empirical linezolid dosing in critically ill patients, in whom pronounced pharmacokinetic variability coincides with increasingly unfavourable MIC distributions. Under these conditions, standard dosing frequently fails to achieve adequate target exposure, and routine TDM as implemented in clinical practice may provide only limited improvement. Systematic model-informed dose individualisation may offer a promising strategy to improve both efficacy and safety. Prospective controlled studies are needed to determine whether MIPD can translate this potential into clinically meaningful benefit.

## Data Availability

UL has full access to the dataset, which is available under request. All R scripts are available in the OSF repository (https://osf.io/n8xd4/).

## 7.3 Author contributions

JS and UL conceptualized the work and developed the methodology. JJ, MZ, CS, JZ, MP, UL were involved in clinical data acquisition. JS, AL performed the model reconstruction. JS, MS analysed, curated and visualised the data. JS, and UL prepared the original manuscript draft. All authors participated in critically revisiting the manuscript. All authors approved the version to be published and agree to be accountable for all aspects of the work. The authors declare that all data were generated in-house, that no paper mill was used and that no AI tool has been used for the generation of text or figures.

### 7.4 Ethics approval and consent to participate

The present study was a secondary analysis of anonymized data from two previously conducted prospective observational studies. No additional ethics approval or consent was required for this re-analysis following consultation with the local ethics committee and in accordance with applicable data-protection regulations (reference: 22-0286 KB). Both original studies had been approved by the Ethics Committee of the Faculty of Medicine, LMU Munich (study 1: 428-12; study 2: 18-578), were conducted in accordance with the Declaration of Helsinki, and obtained written informed consent from all participants.

### 7.5 Clinical trial registration

The present study is a secondary analysis of previously collected data and did not prospectively assign participants to a health-related intervention. Clinical trial number: not applicable. The source observational studies were registered at ClinicalTrials.gov as NCT01793012 on 24 January 2013 and NCT03985605 on 14 June 2019.

## 7.7 Acknowledgements

The authors acknowledge a conference poster entitled “Ten years of susceptibility testing of linezolid in a tertiary care hospital: model-informed precision dosing as a strategy for rising resistance” that was presented at ESCMID Global (Abstr. 00374) in Vienna (11-15 Apr. 2025). A preprint version of this manuscript is available on medRxiv (doi: 10.64898/2025.11.28.25340407).

## 7.8 Declaration of Generative AI and AI-assisted technologies in the writing process

During the preparation of this work the author used ChatGPT (https://chatgpt.com/) and DeepL write (https://www.deepl.com/en/write) in order to improve readability and language of the publication. After using this service, the author reviewed and edited the content as needed and takes full responsibility for the content of the publication.

