## Supplementary material for "Rising resistance to linezolid among critically ill patients calls for model-informed precision dosing in the empirical setting"

PREPRINT

### 1 Methods

#### 1.1 NONMEM® control stream

```
$PROBLEM LZD_Minichmayr_s_2017

$INPUT
ID OID SITE TIME TAD EVID DV BLQ MDV0=MDV MDVAP=DROP MDV1=DROP MDV2=DROP AMT
CMT RATE DUR DI ROUTE OCC COUNTDV LASTDV SCR RRT QUICK BILI AST ALT ALBU LDH THRO
MAP LAC ARDS HB PERITONITIS FIB APACHE WBC SEPSIS CRP ECMO AGE SEX HT TBW

$DATA;
nm_input_sim.csv IGNORE=@;

$SUBROUTINES ADVAN13 TOL6

$MODEL
COMP=(ORAL)
COMP=(CENT)
COMP=(PERI)
COMP=(INHI)
COMP=(AUC)
COMP=(MIC_05)
COMP=(MIC_1)
COMP=(MIC_2)
COMP=(MIC_4)

$PK
iTBW=TBW
IF(TBW.EQ.-99) THEN
iTBW=72.3
ENDIF

iAGE=AGE
IF(AGE.EQ.-99) THEN
iAGE= 67
ENDIF

IF(SCR.LT.0) THEN
CLCR= 63.6
ELSE
CLCR= ((140 - iAGE) / SCR) * (iTBW / 72) * 0.85**SEX
ENDIF

CL = THETA(1)*(1+THETA(9)*(CLCR-80)) *EXP(ETA(1))
V2 = THETA(2)*EXP(ETA(3))
V3 = THETA(3)*EXP(ETA(2))* (iTBW/69.5)
Q = THETA(4)*(iTBW/69.5)**0.75
KA = THETA(5)*EXP(ETA(5))
```

PREPRINT

```
RCLF= EXP(LOG (THETA(6)/(1-THETA(6))) + ETA(4)) / (1+ EXP(LOG (THETA(6)/(1-THETA(6))) +
ETA(4)))
KI = THETA(7)
IC50= THETA(8)

A_0(1) = 0
A_0(2) = 0
A_0(3) = 0
A_0(4) = 0

$DES
ORAL=A(1)
CENT=A(2)
PERI=A(3)
INHI=A(4)

INH= RCLF + (1 - RCLF)*(1- INHI/(INHI+IC50))

DADT(1)= -KA*A(1)
DADT(2)= KA*A(1) - (Q/V2)*A(2) + (Q/V3)*A(3) - A(2)*(CL/V2)*INH
DADT(3)= A(2)*(Q/V2) - A(3)*(Q/V3)
DADT(4)= KI*((A(2)/V2)-A(4))
DADT(5)= A(2)/V2

;Calculation of pk/pd parameters
CC = A(2)/V2
MIC_half=0
MIC_one=0
MIC_two=0
MIC_four=0

IF(CC.GT.0.5) MIC_half=1
IF(CC.GT.1) MIC_one=1
IF(CC.GT.2) MIC_two=1
IF(CC.GT.4) MIC_four=1

DADT(6)=MIC_half ; time above threshold
DADT(7)=MIC_one ; time above threshold
DADT(8)=MIC_two ; time above threshold
DADT(9)=MIC_four ; time above threshold

$THETA
(0, 11.2) ; 1 CL septic
(0, 22.7) ; 2 V2
(0, 19.9) ; 3 V3
(0, 57.9) ; 4 Q
(0, 1.41) ; 5 KA
(0, 0.513) ; 6 RCLF
(0, 0.0017) ; 7 KI
(0, 0.48) FIX ; 8 IC50
(0, 0.00835) ; 9 CLCR_CL
```

PREPRINT

```
$OMEGA
0.149 ; 1 IIV CL
0.144 ; 2 IIV V3
0.126 ; 3 IIV V2
6.45 ; 4 IIV RCLF
0.872 ; 5 IIV KA

$SIGMA
0.024649 FIX ; proportional rsv

$ERROR
IPRED = A(2)/V2
W = SQRT(SIGMA(1)*IPRED**2)
Y = IPRED + IPRED*EPS(1)
IRES = DV-IPRED
IWRES = IRES/(W+0.00001)

AUC = A(5)
MIC_05= A(6)
MIC_1=A(7)
MIC_2=A(8)
MIC_4=A(9)

$ESTIMATION METHOD=1 INTERACTION MAXEVAL=0 SIG=3 PRINT=1 NOABORT POSTHOC
$TABLE ID TIME TAD EVID CMT MDV OCC DV PRED IPRED SITE LASTDV COUNTDV AUC MIC_05
MIC_1 MIC_2 MIC_4 ONEHEADER NOAPPEND NOPRINT FILE=nm_export_rl
```

##### 1.2 Handling of sex and gender dimensions

In this analysis, we employed a binary (female/male) definition of sex in accordance with the data available from previously published prospective studies, where sex was most likely recorded by the admitting physician. We recognize that a binary framework may not fully capture the complexity of sex and gender diversity. In this analysis no sub-group analysis on the basis of sex or gender was employed.

PREPRINT

#### 2 Results

##### 2.1 Distribution of observed dosing intervals

The distribution of the last TDM sample in the first, second and third observed dosing interval regarding the time since first (time after first dose) and most recent dose (time after last dose) is given in Table S1.

***Tab. A1*** *Median (min–max) sampling times (in hours) for TDM samples drawn in different observed dosing intervals (ODIs) in the study 1 and study 2 studies. For each ODI, n indicates the number of samples*

| ODI | n | Time after first dose [h]* | Time after last dose [h]* |
|---|---|---|---|
| | | Study 1 | |
| 1 | 52 | 17.5 (7.3-56.6) | 11.4 (6.2-17.5) |
| 2 | 52 | 29.2 (13.8-68.7) | 11.9 (5.8-23.6) |
| 3 | 52 | 41.6 (25.7-80.3) | 11.9 (6.2-23.9) |
| | | Study 2 | |
| 1 | 65 | 19.6 (3.4-137.5) | 8.2 (0.4-11.6) |
| 2 | 65 | 44.1 (19.9-161.4) | 9.6 (0.1-39.3) |
| 3 | 65 | 68.2 (43.8-184.8) | 9.4 (0.4-25.5) |

*Median (min – max)

#### 2.2 Handling of dose adjustments in study 2

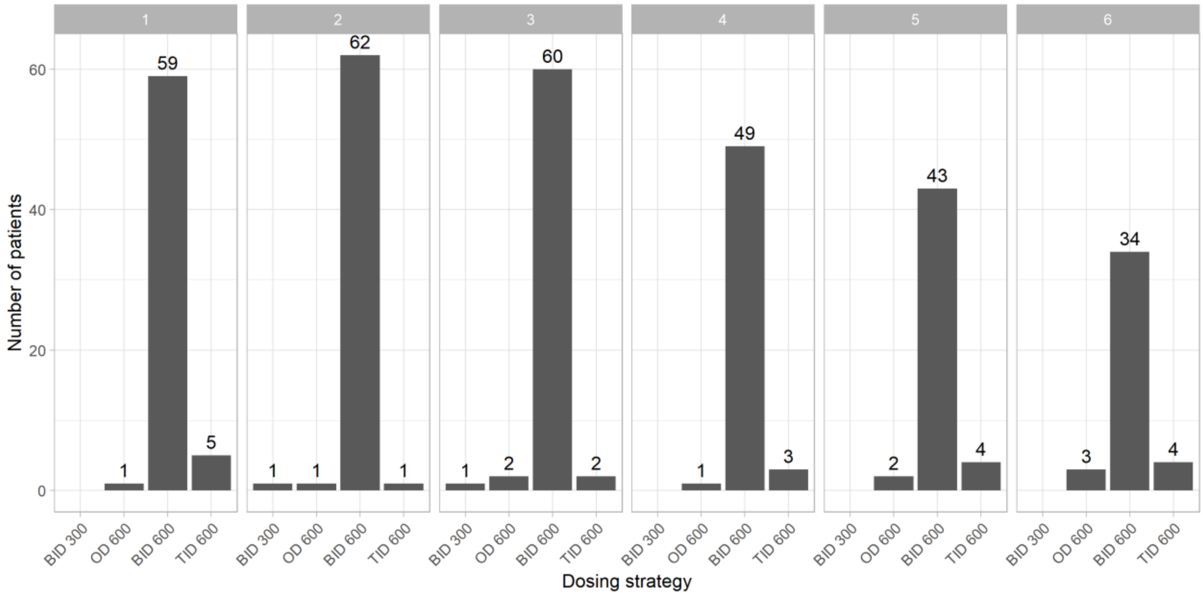


***Fig. A1*** *Distribution of dosing strategies in the TAPSI dataset across observed dosing intervals (panels 1–6). Dosing frequency was classified based on the time span covering three consecutive administrations (i.e., from the previous to the second subsequent dose): three times daily (TID) if the interval was < 30 h, once daily (OD) if > 60 h, and twice daily (BID) if between 30 h and 60 h. This approach distinguishes typical regimens such as 3×8 h (24 h) versus 2×12 h (36 h) dosing. The administered dose levels (300 mg and 600 mg) are shown for each dosing category. Numbers above bars indicate the number of patients in each group.*

Unpublished manuscript version 0.2 (2026-08-06), NOT peer reviewed. Cite at your own risk.

#### 2.3 Cumulative fraction of response per pathogen

***Tab. A2*** *Percentage of patients being under-, overdosed, or reaching empirical target during empirical linezolid therapy for standard dosing, therapeutic drug monitoring, or potential model-informed precision dosing (MIPD). The evaluation is based on pathogen specific minimal inhibitory concentration distributions of pathogens tested for linezolid at intensive care unit in LMU University Hospital.*

| | | | Percentage of patients in category [%] | | | | | | | | |
|---|---|---|---|---|---|---|---|---|---|---|---|
| | | | *S. aureus* | | | Enterococci | | | *S. epidermidis* | | |
| Resistant [%] | | | 0.8 | | | 4.2 | | | 25.5 | | |
| Interval | Study | Regimen | Under | CFR | Over | Under | CFR | Over | Under | CFR | Over |
| ODI1 | Study 1 | Std | 58.3 | 38.9 | 1.9 | 56.3 | 37.6 | 1.8 | 45.0 | 28.0 | 1.4 |
| ODI1 | Study 2 | Std | 25.6 | 65.9 | 7.6 | 26.1 | 62.3 | 7.4 | 21.6 | 47.2 | 5.7 |
| ODI3 | Study 1 | Std | 49.5 | 40.1 | 9.5 | 48.2 | 38.3 | 9.2 | 38.7 | 28.6 | 7.2 |
| ODI3 | Study 2 | TDM | 22.9 | 53.4 | 22.9 | 22.7 | 51.0 | 22.1 | 18.7 | 38.6 | 17.2 |
| ODI3 | Study 1 | MIPD | 3.4 | 91.9 | 3.8 | 6.4 | 85.7 | 3.7 | 5.9 | 65.7 | 2.9 |
| ODI3 | Study 2 | MIPD | 15.1 | 79.5 | 4.6 | 16.5 | 74.9 | 4.4 | 14.2 | 56.8 | 3.4 |

CFR, cumulative fraction of response; MIPD, model-informed precision dosing; ODI1, first observed dosing interval; ODI3, third observed dosing interval; Std, standard; TDM, therapeutic drug monitoring.

PREPRINT
